# Evaluation of efficiency and sensitivity of 1D and 2D sample pooling strategies for SARS-CoV-2 RT-qPCR screening purposes

**DOI:** 10.1101/2020.07.17.20152702

**Authors:** Jasper Verwilt, Jan Hellemans, Tom Sante, Pieter Mestdagh, Jo Vandesompele

## Abstract

To increase the throughput, lower the cost, and save scarce test reagents, laboratories can pool patient samples before SARS-CoV-2 RT-qPCR testing. While different sample pooling methods have been proposed and effectively implemented in some laboratories, no systematic and large-scale evaluations exist using real-life quantitative data gathered throughout the different epidemiological stages. Here, we use anonymous data from 9673 positive cases to simulate and compare 1D and 2D pooling strategies. We show that the optimal choice of pooling method and pool size is an intricate decision with a testing population-dependent efficiency-sensitivity trade-off and present an online tool to provide the reader with custom real-time pooling strategy recommendations.

## Introduction

One of the key strategies in the global battle against the COVID-19 pandemic is massive population testing. However, an ongoing shortage of time, reagents and testing capacity has tempered these efforts. Pooled testing of samples presents itself as a valid strategy to overcome these hurdles and to realize rapid, large-scale testing at lower cost and lower dependence on test reagents.

Multiple recent studies discussed pooling strategies in the frame of SARS-CoV-2 testing. Researchers have explored many strategies, but two of them have been welcomed for their simplicity and effectiveness: one-time pooling (1D pooling) and two-dimensional pooling (2D pooling). In 1D pooling, the samples are pooled, pools are tested and samples in positive pools are tested individually (Figure 1)^1–4^. Labs worldwide have extensively evaluated 1D pooling strategies for SARS-CoV-2 testing in the lab^5–8^ or using simulations^1^. In 2D pooling, samples are organized in a 2D matrix and pools are created along the matrix’s rows and columns. The pools are tested, and negative rows and columns are excluded from the matrix. Next, all remaining samples are tested individually (Figure 1)^9^. Other more complex strategies exist, such as repeated pooling^1^, P-BEST^10^ and Tapestry^11^.

**Figure 1:**
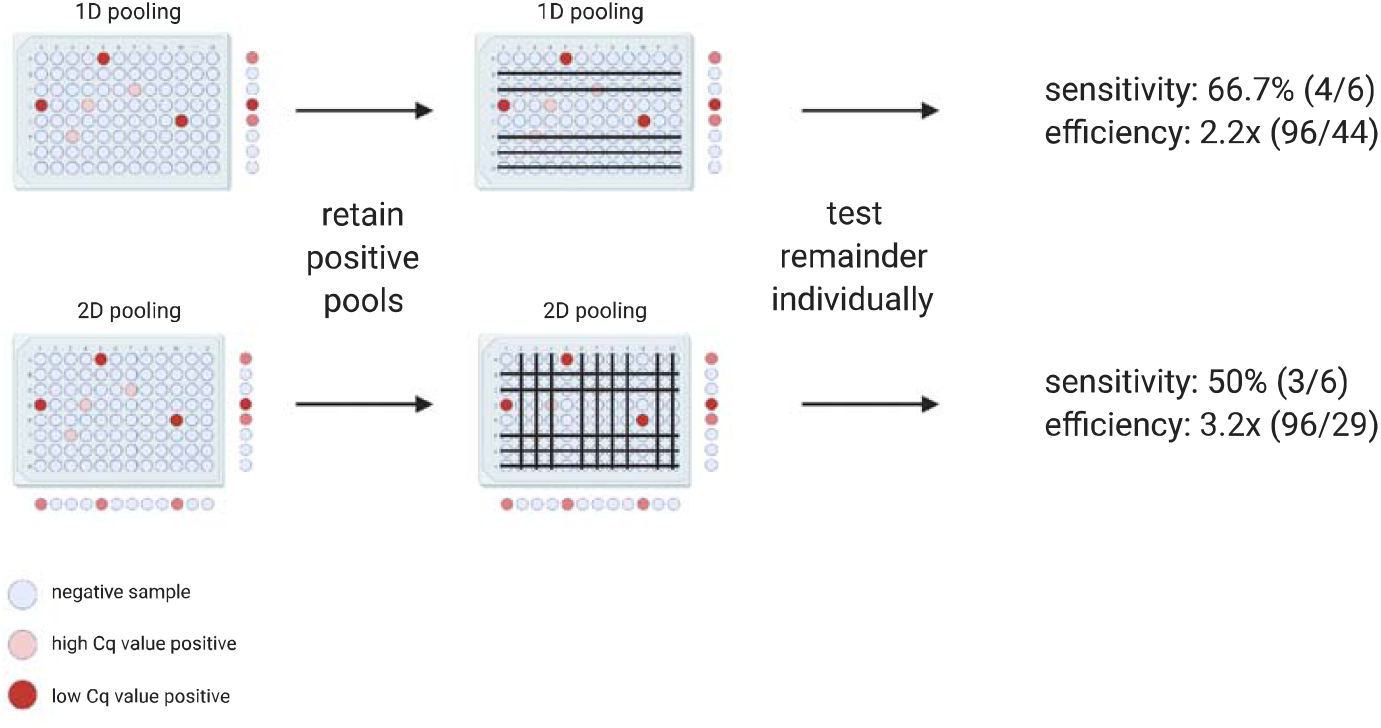
Schematic overview of the applied pooling strategies. The samples are represented as wells in a 96-well microtiter plate. The color of the wells indicates the samples’ SARS-CoV-2 RNA concentration. In 1D pooling, the pools are created by row, the pools are tested and the samples in positive pools are tested again individually. During 2D pooling, the pools are created by row and column (each sample exists in two pools), the pools are tested, all negative rows and columns are removed and the remaining samples are tested individually. The sensitivity and the efficiency are calculated according to the equations found in the methods.

While attractive, pooling strategies come with inherent limitations. First, pooling dilutes each sample, possibly to such an extent that the viral RNA becomes undetectable, which results in false negative observations^8,9,12^. A second limitation is that an increase in sample manipulations augments the risk of cross-contamination and sample mix-ups, possibly leading to false negatives and false positives^9^. Last, when pooling, identifying individual positive samples will take an additional RNA extraction and RT-qPCR run, while one run is sufficient when testing individual samples without pooling, which increases labor and turnaround time.

Although the number of preprints and peer-reviewed publications on pooling strategies for COVID-19 RT-qPCR-based testing has accelerated rapidly throughout the pandemic, some critical aspects remain mostly ignored. First of all, the proposed optimal pooling strategy is most often based on a binary classification of samples as either positive or negative. This Boolean approach is not true to the real-world situation and does not investigate the pooling step’s dilution effect. Second, when using Cq values as a semi-quantitative measure^13^ of the viral loads, their overall distribution should reflect the real-life population. A high fraction of Cq values close to the limit of detection of the RT-qPCR assay produces an elevated risk of resulting in false negative samples^14^. Last, since the Cq distribution of the sample population and prevalence may vary over time, it remains unclear how the pooling strategy’s performance evolves as the pandemic progresses.

We questioned to what extent optimal pooling strategies would have changed throughout the COVID-19 pandemic and how testing facilities might use pooling strategies for future testing in a correct and attainable manner. To this extent, we simulated and evaluated one-dimensional (1D) and two-dimensional (2D) pooling strategies with different pool sizes using real-life RT-qPCR data gathered by the Belgian national testing platform during the end of the first and the beginning of the second SARS-CoV-2 epidemiological waves. Additionally, we formulate a detailed action plan to provide testing laboratories with the most suitable pooling strategy assuring an optimal efficiency-sensitivity trade-off.

## Materials and Methods

### Patient samples

Nasopharyngeal swabs were collected in VTM or DNA/RNA Shield (Zymo Research) by a healthcare professional as a diagnostic test for SARS-CoV-2, as part of the Belgian national testing platform. The individuals were tested at nursing homes or in triage centers, between April 9^th^ and June 7^th^, and between September 1^st^ and November 10^th^. After filtering the data as described further, this resulted in 207 944 patients in total, of which 9673 positives (4.65%).

### SARS-CoV-2 RT-qPCR test

During the first (spring) wave, RNA extraction was performed using the Total RNA Purification Kit (Norgen Biotek #24300) according to the manufacturer’s instructions using 200 µl transport medium, 200 µl lysis buffer and 200 µl ethanol, with processing using a centrifuge (5810R with rotor A-4-81, both from Eppendorf). RNA was eluted from the plates using 50 µl elution buffer (nuclease-free water), resulting in approximately 45 µl eluate. RNA extractions were simultaneously performed for 94 patient samples and 2 negative controls (nuclease-free water). After addition of the lysis buffer, 4 µl of a proprietary 700 nucleotides spike-in control RNA (prior to May 25^th^, 40 000 copies for singleplex RT-qPCR; from May 25^th^ onwards, 5000 copies for duplex RT-qPCR) and carrier RNA (200 ng of yeast tRNA, Roche #10109517001) was added to all 96 wells from the plate. To the eluate of one of the negative control wells, 7500 RNA copies of positive control RNA (Synthetic SARS-CoV-2 RNA Control 2, Twist Biosciences #102024) were added. During the second (autumn) wave, RNA extraction was performed using the Quick-RNA Viral 96 Kit (Zymo Research #R1041), according to the manufacturer’s instructions using 100 µl transport medium, with processing using a centrifuge (5810R with rotor A-4-81, both from Eppendorf). RNA was eluted from the plates using 30 µl elution buffer (nuclease-free water). RNA extractions were simultaneously performed for 92 patient samples, 2 negative controls (nuclease-free water), and 2 positive controls (1 diluted positive case as a full workflow control; 1 positive control RNA as RT-qPCR control, see further). After addition of the lysis buffer, 4 µl of a proprietary 700 nucleotides spike-in control RNA (5000 copies) and carrier RNA (200 ng of yeast tRNA, Roche #10109517001) was added to all 96 wells from the plate. To the eluate of one of the negative control wells, 7500 RNA copies of positive control RNA (Synthetic SARS-CoV-2 RNA Control 2, Twist Biosciences #102024) were added.

Six µl of RNA eluate was used as input for a 20 µl RT-qPCR reaction in a CFX384 qPCR instrument using 10 µl iTaq one-step RT-qPCR mastermix (Bio-Rad #1725141) according to the manufacturer’s instructions, using 250 nM final concentration of primers and 400 nM of hydrolysis probe. Primers and probes were synthesized by Integrated DNA Technologies using clean-room GMP production. For detection of the SARS-CoV-2 virus, the Charité *E* gene assay was used (FAM)^15^; for the internal control, a proprietary hydrolysis probe assay (HEX) was used. Prior to May 25th, 2 singleplex assays were performed; from May 25^th^ onwards, 1 duplex RT-qPCR was performed. Cq values were generated using the FastFinder software v3.300.5 (UgenTec). Only batches were approved with a clean negative control and a positive control in the expected range.

### Digital PCR calibration of positive control RNA

Digital PCR was done on a QX200 instrument (Bio-Rad) using the One-Step RT-ddPCR Advanced Kit for Probes (Bio-Rad #1864022) according to the manufacturer’s instructions. Briefly, 22 µl pre-reactions were prepared consisting of 5 µl 4x supermix, 2 µl reverse transcriptase, 6 µl positive control RNA (125 RNA copies/µl), 15 mM dithiothreitol, 900 nM of each forward and reverse primer and 250 nM *E* gene hydrolysis probe (FAM) (see higher). 20 µl of the pre-reaction was used for droplet generation using the QX200 Droplet Generator, followed by careful transfer to a 96-well PCR plate for thermocycling: 60 min 46 °C reverse transcription, 10 min 95 °C enzyme activation, 40 cycles of 30 sec denaturation at 95 °C and 1 min annealing/extension at 59 °C, and finally 10 min 98 °C enzyme deactivation. Droplets were analyzed by the QX200 Droplet Reader and QuantaSoft software. With an RNA input of 7500 copies per reaction, the digital PCR result was 1500 cDNA copies (or 20% of the expected number, a fraction confirmed by Dr. Jim Huggett for particular lot numbers of #102024, personal communication. The reason for this discrepancy is two-fold: the number of RNA molecules provided by the manufacturer is only approximate, and the reverse transcription reaction is inefficient). The median Cq value of the positive control RNA of 24.55 thus corresponds to 1500 digital PCR calibrated cDNA molecules.

### Determination of efficiency and sensitivity for simulated of 1D and 2D pooling strategies

Simulations are run using R 4.0.1. First, several cohorts of 100 000 patients are repeatedly simulated with varying fractions of positive cases ***f***, depending on the Simulations are run using R 4.0.1. First, several cohorts of 100 000 patients are fraction of positive samples of the investigated week. This is done five times, resulting in five replicate cohorts per week. The Cq values of the positive samples are sampled with replacement from the set of the positive Cq values of said week. Next, the patients are randomly separated into pools depending on the pooling strategy that is simulated. The pooling strategies that were simulated are 1×4, 1×8, 1×12, 1×16, 1×24 (all 1D), and 8×12, 12×16 and 16×24. The Cq value of the pool was calculated as follows:

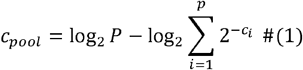

With *C*_*pool*_ the Cq value of the pool, *P* the number of samples in the pool, *p* the number of positive samples in the pool, *C*_1_, *C*_2_, …, *C*_*p*_ the Cq values of the positive samples. If the Cq value of the pool is lower than the single-molecule Cq value, it is classified as a positive pool. For 1D pooling, only samples in positive pools are retained and the remaining individual Cq values were checked to be positive. For 2D pooling, the Cq values of the differently sized pools are checked simultaneously and the samples in negative pools are removed, after which all Cq values of the remaining samples are checked individually. Samples that were retained after the testing of the pools and that had an individual Cq lower than the single-molecule Cq value are classified as positive, all other samples are classified as negative.

The sensitivity is calculated as:

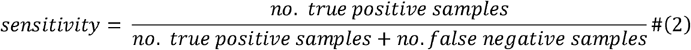

The analytical efficiency gain is calculated as:

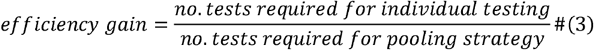

In all simulations, the number of tests required for individual testing is equal to the number of samples (assuming no technical failures). The outcomes for each simulation were identical as the sample size far outreached the size of the dataset. The code is available at https://github.com/OncoRNALab/covidpooling.

### Ad hoc sensitivity and efficiency calculation

To calculate the efficiency for a specific 1D pooling strategy on a real sample set, the following equation was used:

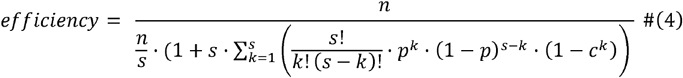

With sample size *n*, pool size *s*, fraction of positive samples *p* and fraction of Cq values of positive samples above the ‘dilution detection limit’: the lowest individual Cq value that can result in a pooled Cq value lower than the single molecule Cq value, or:

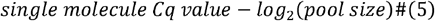

Equation (4) is derived as follows. The efficiency is defined by the following equation:

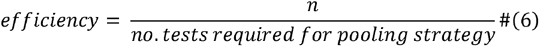

The number of tests performed when using a pooling strategy is equal to:

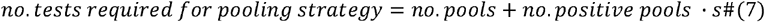

Since 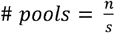,

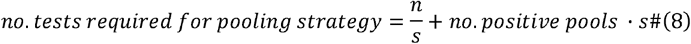

The exact number of positive pools can be calculated by multiplying the number of pools by the probability of a pool testing positive. Approximately, a pool will test positive if it includes a positive sample with a Cq value lower than the ‘dilution detection limit’. The probability of having a specific number of positive samples *k* in a pool with pool size *s* is defined by a binomial distribution:

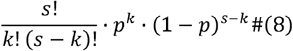

Thus, the probability of having at least one positive value in a pool is equal to:

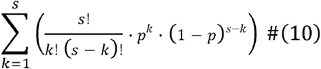

In general, we can assume that when a sample has a Cq value higher than the ‘dilution detection limit’, for the sample to test positive, it must be accompanied by a sample with a Cq value lower than the ‘dilution detection limit’. Equation (10) can be adjusted to factor for these events:

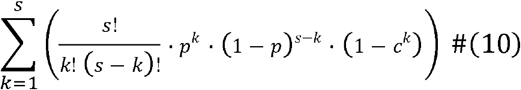

Filling in Eq. (10) in Eq. (8) results in the final formula being used for the calculation of the efficiency.

To estimate the sensitivity for a specific 1D pooling strategy on a real sample set, the following equation was used:

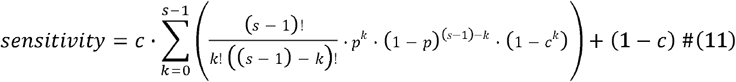

The sensitivity can be defined as the probability a true positive sample tests positive. For our situation it will be equal to the probability that any sample tests positive:

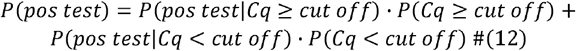

Previously, *P*(*Cq ≥ cut off*) was defined as *c* and therefore *P*(*Cq ≥ cut off*) = 1 – *c*. Also *P*(*pos test* |*Cq < cut off*) = 1. A positive sample with Cq value above the ‘dilution detection limit’ can only test positive if one of the other samples in the pool is also positive and has a Cq value lower than the ‘dilution detection limit’. We can calculate the probability of this happening by using the same logic as before, but with *s* – 1 instead of *s:*

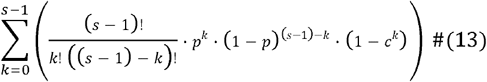

Completing Eq. (12) with Eq. (13) leads to Eq. (11) for calculating the sensitivity.

### Web application

To help laboratories find the best pooling strategy for their specific situation (i.e. the local positivity ratio and Cq value distribution), we developed a Shiny application in R 4.0.1. The Shiny application was launched on our in-house Shiny server and is available at https://shiny.dev.cmgg.be/.

## Results

### Single-molecule Cq value determination

We made a 5-point 10-fold serial dilution series of positive control RNA from 150 000 (digital PCR calibrated) copies down to 15 copies. The Y-intercept value points at a single-molecule Cq value of 35.66 and 35.28 for singleplex and duplex RT-qPCR, respectively (Supplemental Figure 1). Therefore, we conservatively use 37 as the single-molecule value for further analysis. Patient sample Cq values higher than the single-molecule Cq value threshold are likely due to random measurement variation, lot reagent variability and sample inhibition.

### Cq distribution is dynamic over course of the pandemic

Few studies have explored how the Cq value distribution within one testing facility evolves during the COVID-19 pandemic. We determined the 75%-tile of the Cq value distribution and the percentage of positive tests per day as a proxy for actual Cq value distribution and prevalence, respectively (Figure 2). We compared the fraction of positive tests in our dataset with the fraction of positive tests as reported by the federal agency for public health Sciensano (https://epistat.wiv-isp.be/covid/. accessed January 25^th^, 2021). First, the fractions of positive tests seem to align at the end of the first wave, but in the second wave our data seems to be shifted about one to two weeks later. Second, the 75%-tile of the Cq values varies over the course of the pandemic from a minimum value of around 18 and a maximum value of almost 35. Third, when comparing the fraction of positive samples and the 75%-tile of the Cq value distribution, we note that these parameters are inversely related: when the positivity rate goes down, the Cq value distribution shifts towards the higher end of the spectrum (Supplemental Figure 2). In conclusion, the Cq value distribution and prevalence show a dynamic profile over the course of the COVID-19 pandemic. These observations are crucial considering that positivity rate and Cq value distribution are key determinants of efficiency and sensitivity of any pooling strategy.

**Figure 2:**
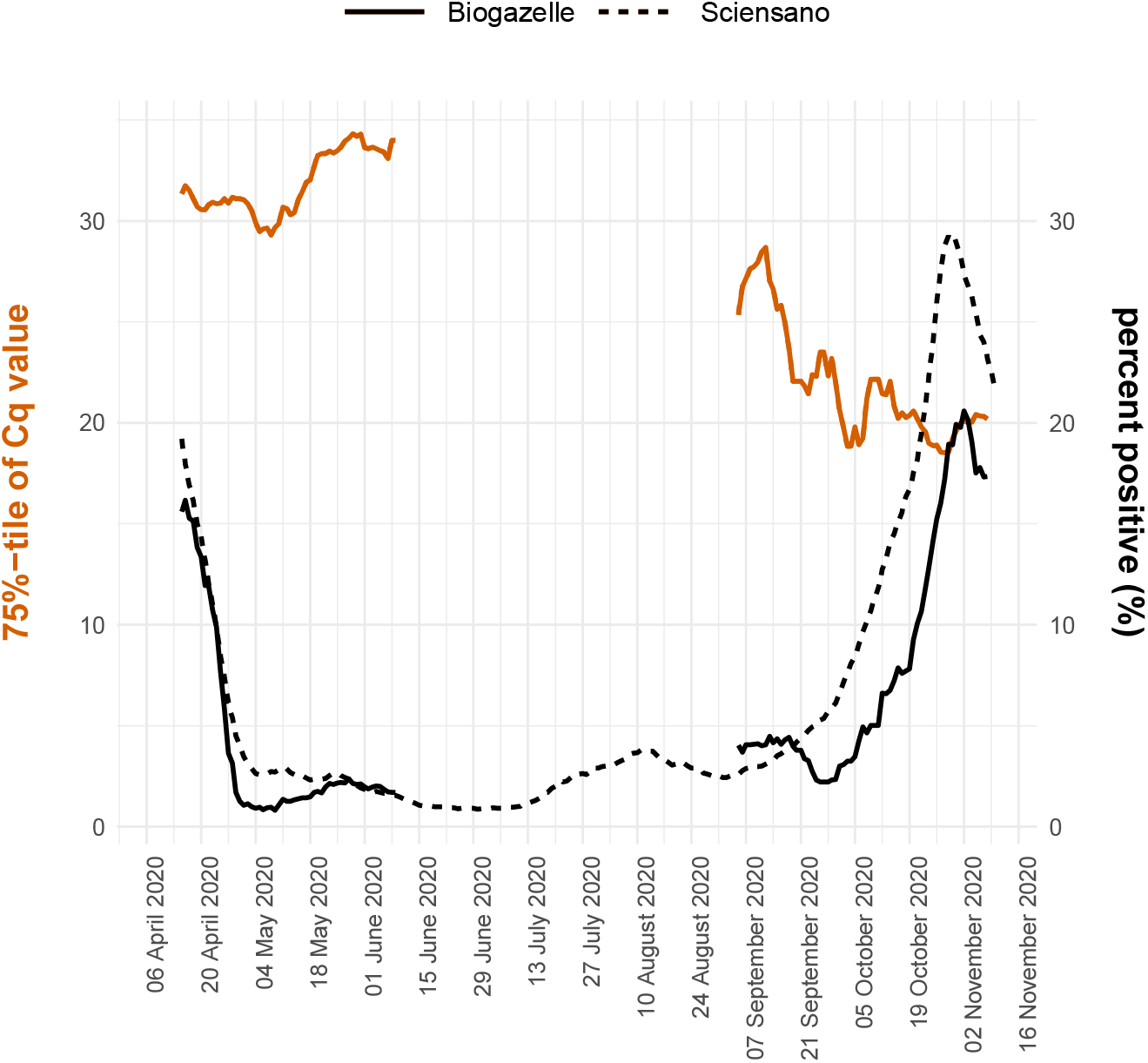
Evolution of the 75%-tile of the Cq value distribution and fraction of positive samples. The left y-axis shows the seven day moving window average of the 75%-tile of the Cq value distribution of the data originating from Biogazelle and the right y-axis shows the seven day moving window average of the fraction of positive samples for the Biogazelle and Sciensano data. The two datasets are differentiated by the line type. If the moving average was calculated using on the basis of less than five days (due to no data being available for specific days), the datapoint was removed from the visualization.

### Pooling efficiency and sensitivity changes as pandemic progresses

To explore how hypothetical pooling strategies would have affected the SARS-CoV-2 testing outcomes, we simulated different 1D (with pool size of 4, 8, 12, 16 and 24) and 2D pooling (with pool sizes of 8×12, 12×16, and 16×24) strategies using individual sample Cq values from a single Belgian laboratory during the end of the first and beginning of the second wave. The data was grouped by week and the resulting Cq value distributions and positivity rates were used as input for the simulations (Figure 3). First, sensitivity and efficiency show very opposing patterns when comparing different timeframes during the pandemic. At the end of the first wave the efficiency increases, while at the beginning of the second wave, the efficiency decreases. The sensitivity drops as we move further away from the first wave but remains stable as we enter the second. Second, pool size and strategy have a major influence on the outcomes. 2D pooling strategies generally have the highest efficiency, but the lowest sensitivity. Curiously, strategies with larger pool sizes were more efficient during the end of the first wave, but less efficient during the beginning of the second wave. The sensitivity was always higher for strategies with smaller pool sizes, irrespective of the time during the pandemic. We conclude that— just like the positivity rate and the Cq value distribution—the sensitivity and efficiency depend on the timing in the pandemic and are heavily affected by the pooling strategy and the size of the pools.

**Figure 3:**
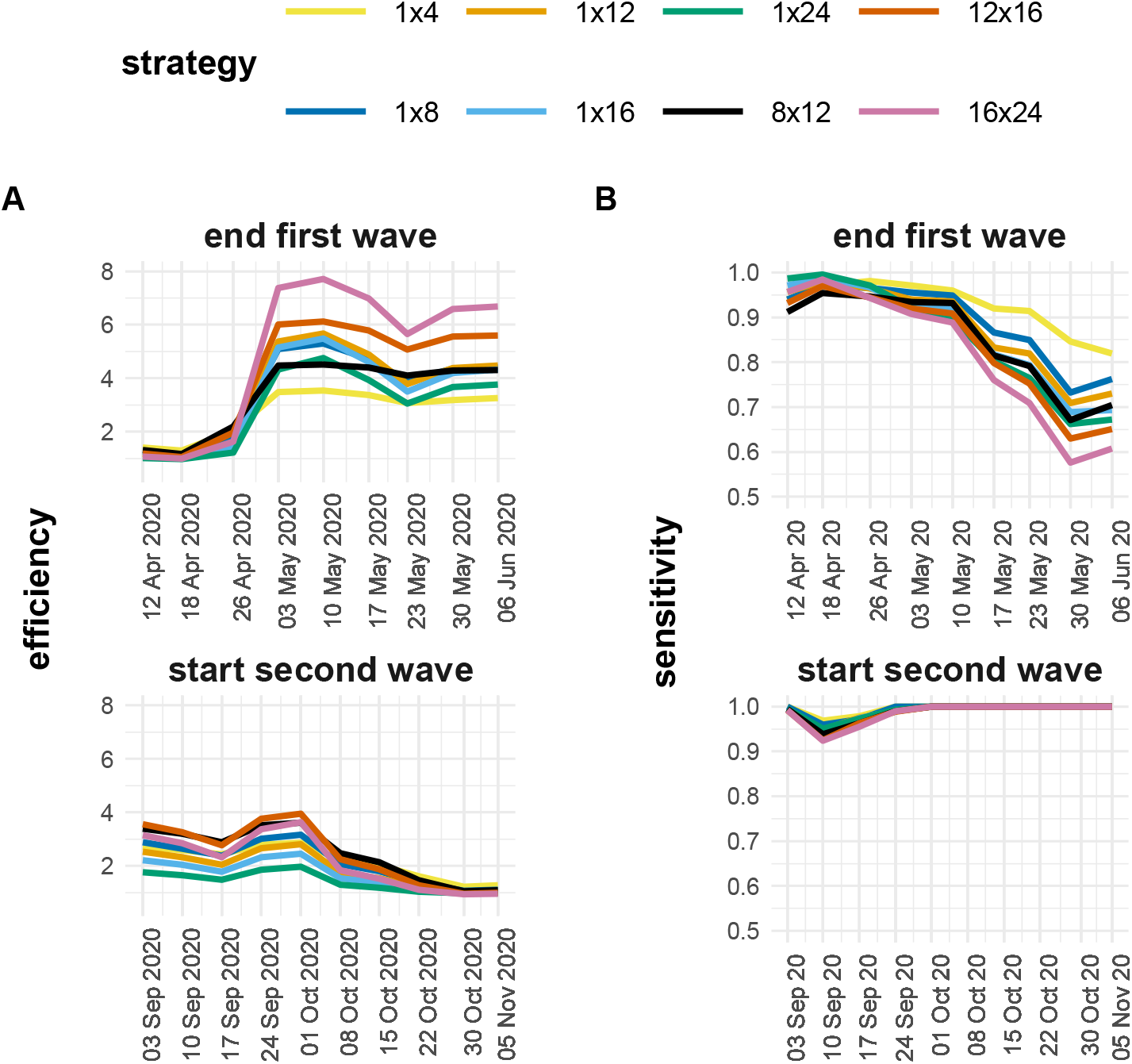
Sensitivity and efficiency for the end of the first (A) and the start of the second (B) Belgian SARS-CoV-2 infection wave. The data is grouped by week and the sensitivity and efficiency are calculated by simulating different pooling strategies (1×4, 1×8, 1×12, 1×16, 1×24, 8×12, 12×16 and 16×24). The pooling strategies can be distinguished by color.

### Positivity rate drives efficiency, Cq distribution drives sensitivity

We wondered how the positivity rate, Cq value distribution and pooling strategy affect the performance of the adopted strategy. To investigate this, we used the previous simulations for the end of the first wave to create an adjusted visualization where all parameters involved are incorporated (Figure 4). First, it is apparent that weeks with a high 75%-tile Cq value tend to have a low sensitivity and weeks with a high positivity rate seem to have a low efficiency. Second, pooling strategies with smaller pool sizes seem less sensitive to changes in positivity rate and Cq value distribution, as indicated by the area of the polygon traced around the edges of the data (Figure 4). These results show that the prevalence mainly contributes to the efficiency and the Cq distribution to the sensitivity.

**Figure 4:**
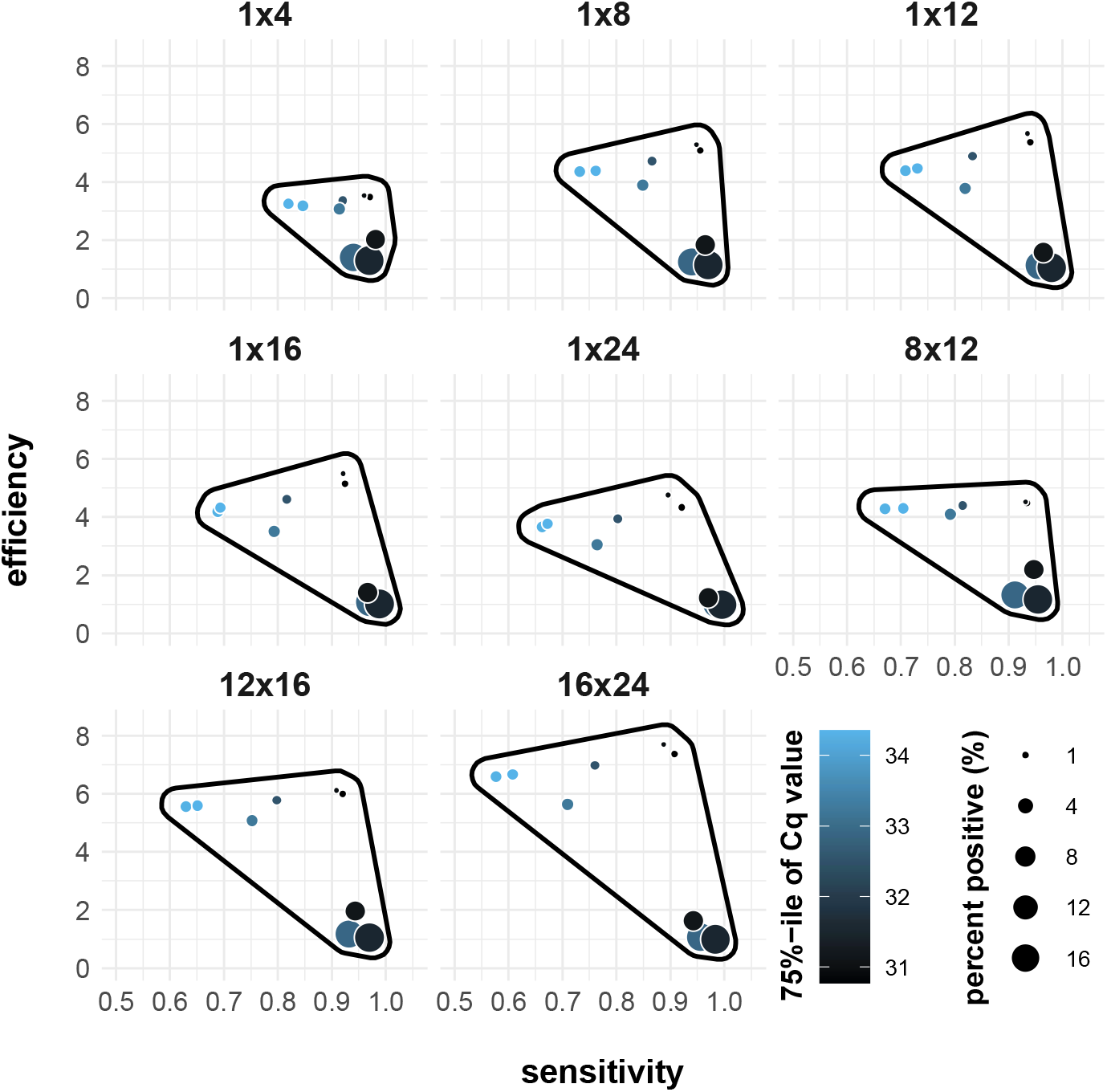
Simulated sensitivity and efficiency for the end of the first wave visualized with relation to the week (different circles), fraction of positive samples (size of circles) and 75%-tile of the Cq value distribution (color). A polygon is drawn around the datapoints (with a small margin) to visualize and to compare the variability of the sensitivity and efficiency over a period of time between pooling strategies.

### Shiny app for guided decision making

To provide laboratories with a custom pooling strategy recommendation based on their specific sampling population, we worked out equations to estimate the sensitivity and efficiency (for 1D pooling strategies) based on an uploaded dataset of Cq values. The derivation of these equations can be found in the Methods section. We focused on 1D pooling strategies since 2D pooling strategies generally resulted in extreme outcomes (highest efficiency and lowest sensitivity) and the outcomes of the optimal pooling strategy are situated somewhere in two extremes. To evaluate the equations’ capacities to replicate the simulations, we compared the simulated efficiency and sensitivity of the pooling strategies for the different weeks and the efficiency and sensitivity of the pooling strategies the distributions, fraction of positive samples and single-molecule cutoff as inputs for the formulas (Supplemental Figure 3 and Supplemental Figure 4). We integrated these formulas into an open-access Shiny application (Supplemental Figure 5). The application requires three inputs: a dataset of Cq values from positive samples, the positivity rate and the single-molecule cut-off Cq value. The Shiny application will then swiftly output the estimated data-specific efficiency and sensitivity for different pooling strategies.

## Discussion

Using a sizeable real-life dataset of 9673 SARS-CoV-2 positive nasopharyngeal samples, we found that the pooling strategies’ sensitivity and efficiency mainly depend on the prevalence and the distribution of the Cq values. Our results indicate that both the prevalence and the Cq value distribution are dynamic parameters during the SARS-CoV-2 pandemic and that, as a result, the resulting sensitivity and efficiency of pooling strategies are as well. To enable researchers and institutions with a real-time and accessible recommendation concerning the optimal 1D pooling strategy for their testing population, we developed a Shiny app providing just that. Two factors could explain the dynamics of the prevalence and the Cq value distribution: epidemiological and virological change within the same sampling population and variation in the sampling population. The existence of these factors would suggest that an intricate interplay of these two components is at the origin of the observed evolutions. Recent research indicated that the first component (epidemiological change) exists, as the distribution of random surveillance testing-deduced Cq values fluctuates during the SARS-CoV-2 pandemic (by definition, no changes in sampling population occurred in this research, thereby excluding this factor from the equation)^16^. As such, an alarming change in reproduction number (inherently coupled to the epidemiological period) should induce a reassessment of the pooling strategy^17^. The second component (variation in sampling population) is bound to happen when the testing facility is not consistently receiving samples from the same origin, as is the case for Biogazelle. At the very introduction of Biogazelle as a testing facility, most samples originated from hospitals and sources were added progressively as the testing capacity increased. Additionally, the Belgian government instituted a rapid change in the testing regime on October 21^st^, 2020: only symptomatic suspected SARS-CoV-2 cases get tested. The federal government lifted this measure on November 23^rd^, 2020, when the number of cases lowered and the existing testing capacity sufficed again. Since symptomatic patients generally show lower Cq values^18,19^, it is clear that sampling bias will contribute to the overall Cq value distribution.

The influence these dynamic parameters have on the variation of performance of pooling strategies is significant. This observation raises an issue for interpreting pooling strategy evaluations not based on time-series datasets. The effectiveness of a chosen pooling plan might even decrease to such an extent that it becomes inferior to individual testing. We observed this situation at the end of the second wave when efficiency is close to 1, but sensitivity is not (Figure 3). Based on these results, it becomes essential to regularly re-evaluate an adopted pooling strategy to avoid compromising on sensitivity and efficiency when there is no need.

Multiple effects contribute to how the testing population’s characteristics drive pooling strategy outcomes. The main trends show that the prevalence mainly influences efficiency, and the Cq value distribution mainly influences sensitivity (Figure 3). We can explain both observations by using common sense and basic mathematics. When the prevalence is low, the efficiency is high: fewer pools will have positive samples and therefore test negative, which will automatically result in a lower number of tests needed to test all samples. Additionally, when a considerable proportion of samples have a Cq value close to the single-molecule Cq value, a more significant fraction of samples will become too diluted to detect during pooling and result in false negatives. There appear to be secondary compensating effects of the Cq value distribution and prevalence on the efficiency and sensitivity, respectively, which are more subtle. Primarily, as a higher fraction of positive samples has a Cq value close to the upper limit, more pools will test (false) negative, boosting the efficiency. On the other hand, when the prevalence increases, the sensitivity will increase due to an effect we call ‘rescuing’: a high Cq value that would otherwise test negative when diluted in the pool is ‘rescued’ by a low Cq value in the same pool. When the prevalence rises, the chances of this phenomenon happening also increase and as will the sensitivity. The same was observed by Cleary et al.^17^. Although minor, these secondary effects explain a number of our observations.

To elaborate how the optimal pooling strategy (best efficiency trade-off) transforms over time, assume two situations: low prevalence and high prevalence. When the prevalence is low, the larger pool sizes will result in higher efficiency and lower prevalence (more dilution). However, when the prevalence is high, the ‘rescuing’ effect will be more prominent and counteract the increasing efficiency and decreasing sensitivity. These results are in line with the widely accepted idea that sample pooling methods show a higher efficiency when pool size is large and that as prevalence increases, it reached a threshold after which smaller pool sizes become more efficient^1,9^. Intuitively, the ‘rescuing’ effect is less prominent in 2D pooling strategies, as both pools (row and column) need to rescue the high Cq sample.

False negatives have pre-pool Cq values close to the detection limit and predominantly originate from patients who are at the end of an infection^17,20^, putting their clinical relevance in question (i.e. no longer infectious). Similarly, however, one can argue that these high Cq samples are imperative to a favorable pandemic response: they might originate from pre-symptomatic or very recently-infected patients^17^, allowing for catching cases before transmission—a principle at the very core of every population screening strategy. Also, we cannot rule out that these high Cq values are due to imperfect sampling or any other mistakes along the sample preparation^13^.

Our study suffers from some essential limitations. First, although the data grouped by weeks provides many different situations to assess, there will still be other combinations of parameters that we did not analyze in this paper. However, the current dataset probably represents the most plausible scenarios as the data originates from a protracted period of the pandemic. Second, we selected only 1D and 2D pooling methods in this simulation study. As stated before, other pooling regimes exist and might be more performant than the discussed ones. Yet, these pooling strategies come with intrinsic shortcomings. The P-BEST pooling protocol is very time consuming^10^, even when using a pipetting robot, and the repeated pooling method suffers from a complicated re-pooling scheme^1^. Third, our model relies on the critical assumption that we can directly induce the pool’s Cq value from the individual samples’ Cq values using a simple formula (see Methods). Wet lab experiments have shown that this is not necessarily the case^5–8^. Fourth, to calculate the pooling strategies’ performance, the single-molecule Cq value and the prevalence must be known. However, we can easily calculate the single-molecule Cq value by generating a ddPCR calibrated dilution series as done in this paper. A testing laboratory can also choose to utilize a cut-off Cq value different from the single-molecule Cq value. Once a threshold Cq value is determined, it should not be changed. The prevalence, however, cannot be known precisely, and as a result, the prevalence must be estimated. We can do this either before adopting a pooling strategy by testing the individual samples and using the fraction of positive samples as an indication for the prevalence or when a pooling strategy is already in place by calculating it from the percentage of positive pools^2,17^. Last, the calculated efficiency gain is merely a representation of the number of individual RNA extractions and RT-qPCR reactions and does not evaluate the amount of labor or time-to-result. Pooling a low number of samples will unnecessarily increase the time-to-result and workload. In conclusion, we show that finding the optimal pooling strategy for SARS-CoV-2 test samples is guided by a testing population-dependent efficiency-sensitivity trade-off. Consequently, the most favorable pooling regime might change throughout the pandemic due to epidemiological changes and revisions in diagnostic testing strategies. We provide an accessible shiny application to guide readers towards the optimal pooling strategy to fit their needs.

## Data Availability

The code and Cq values are available on https://github.com/OncoRNALab/covidpooling.

## Acknowledgements

We are grateful for the data from the Belgian federal taskforce for COVID-19 qPCR testing.

Conceptualization: J.Va., P.M. and J.Ve.; Methodology: J.Va., P.M. and J.Ve.; Software: J.Ve., T.S.; Formal Analysis: J.Ve.; Resources: J.H., J.Va. and P.M.; Data Curation: J.Ve.; Writing – Original Draft: J.Va. and J.Ve.; Writing – Review & Editing; J.Va., J.H., P.M., T.S. and J.Ve.; Visualisation: J.Va. and J.Ve.; Supervision: J.Va and P.M.; Project Administration: J.Va. and P.M.

## Data availability

The code and Cq values are available on https://github.com/OncoRNALab/covidpooling.

---

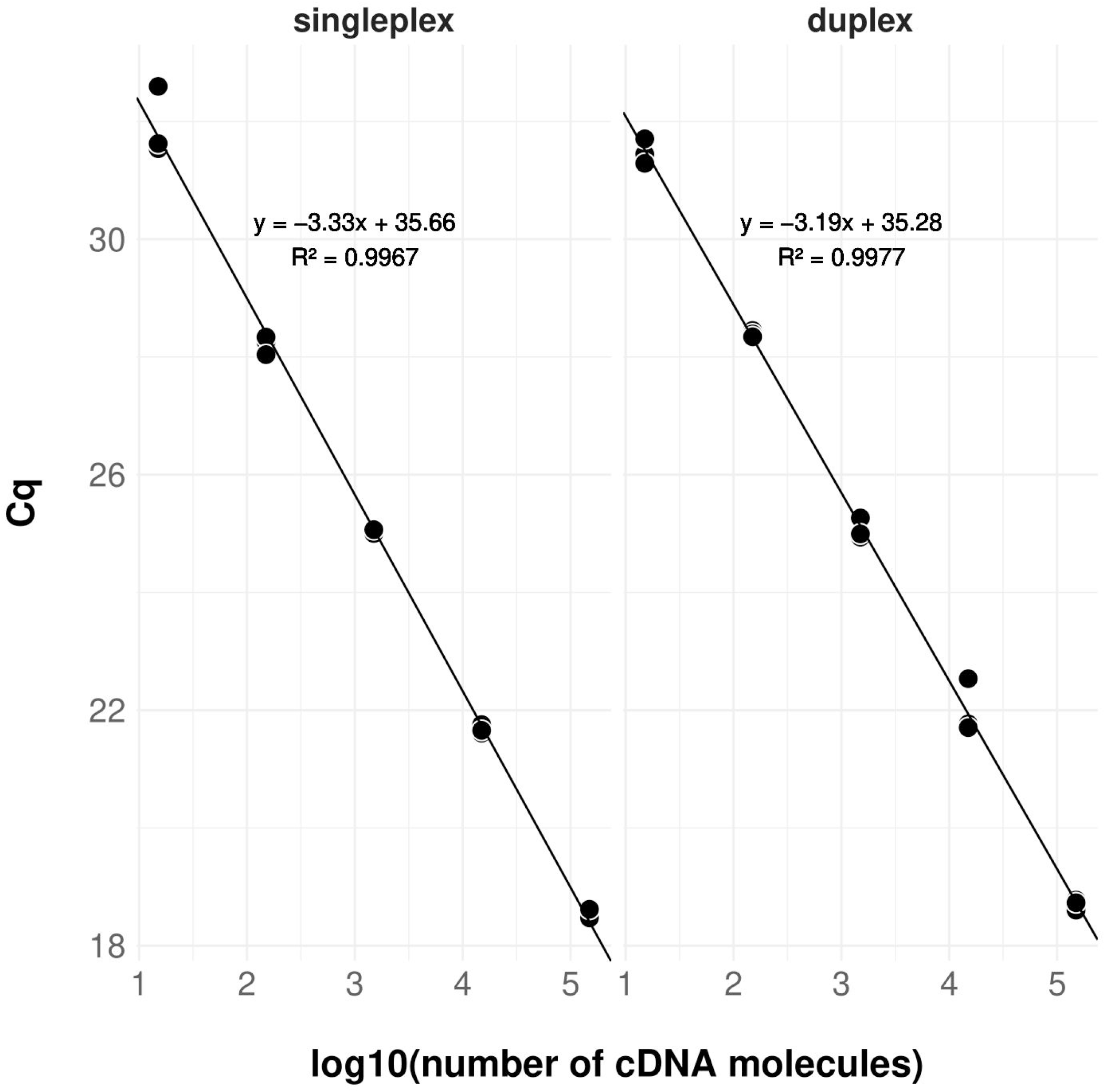

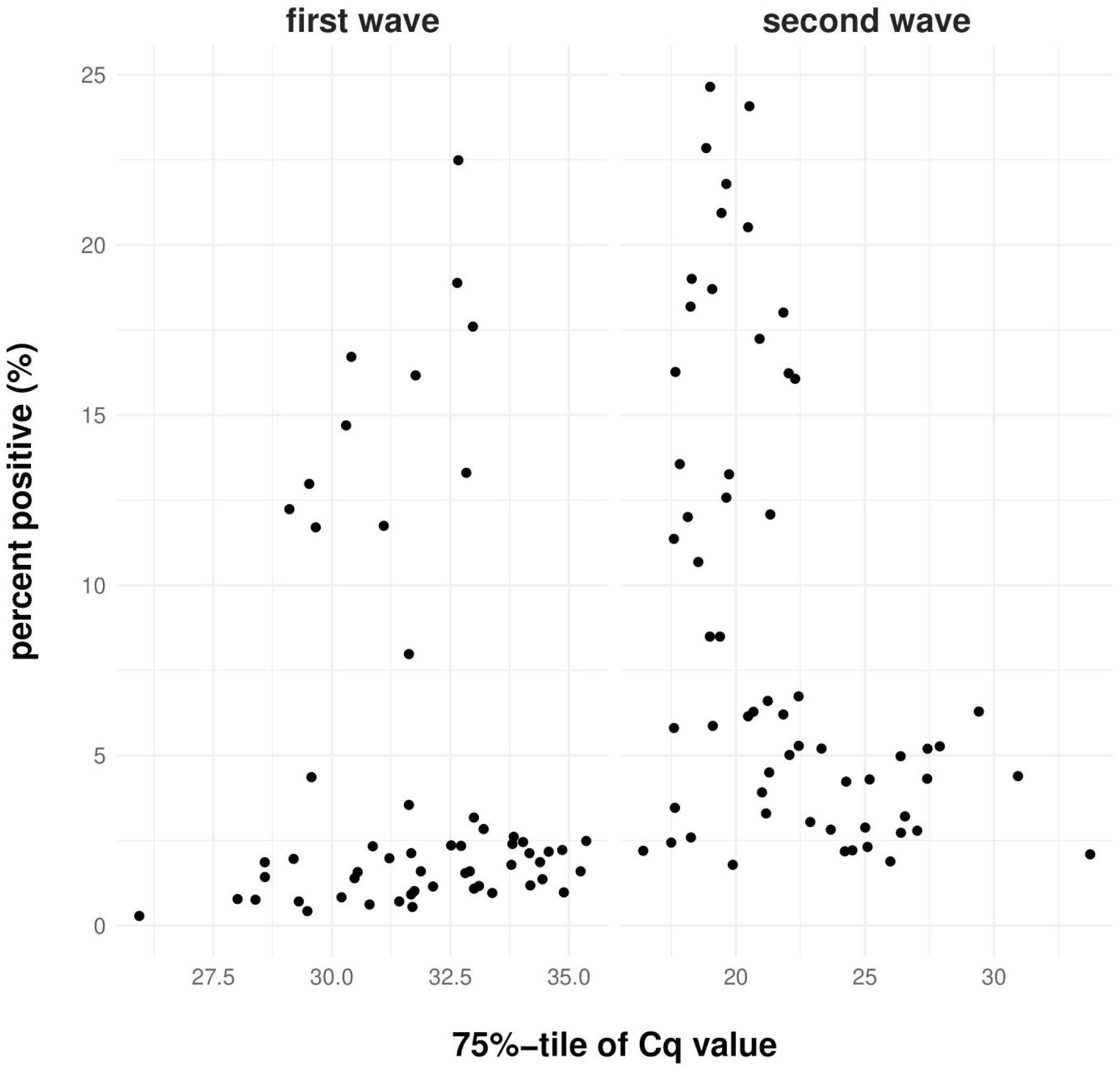

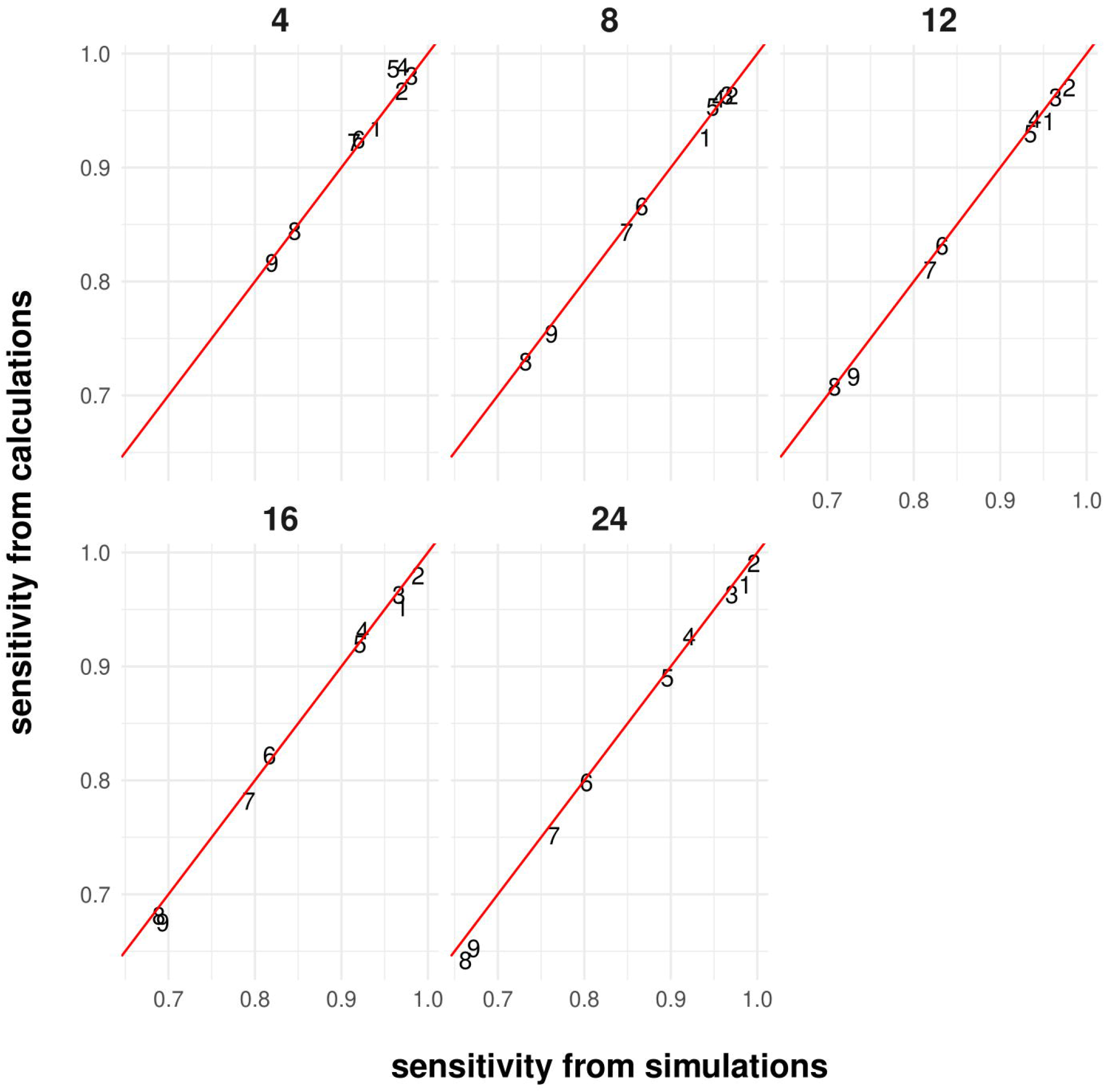

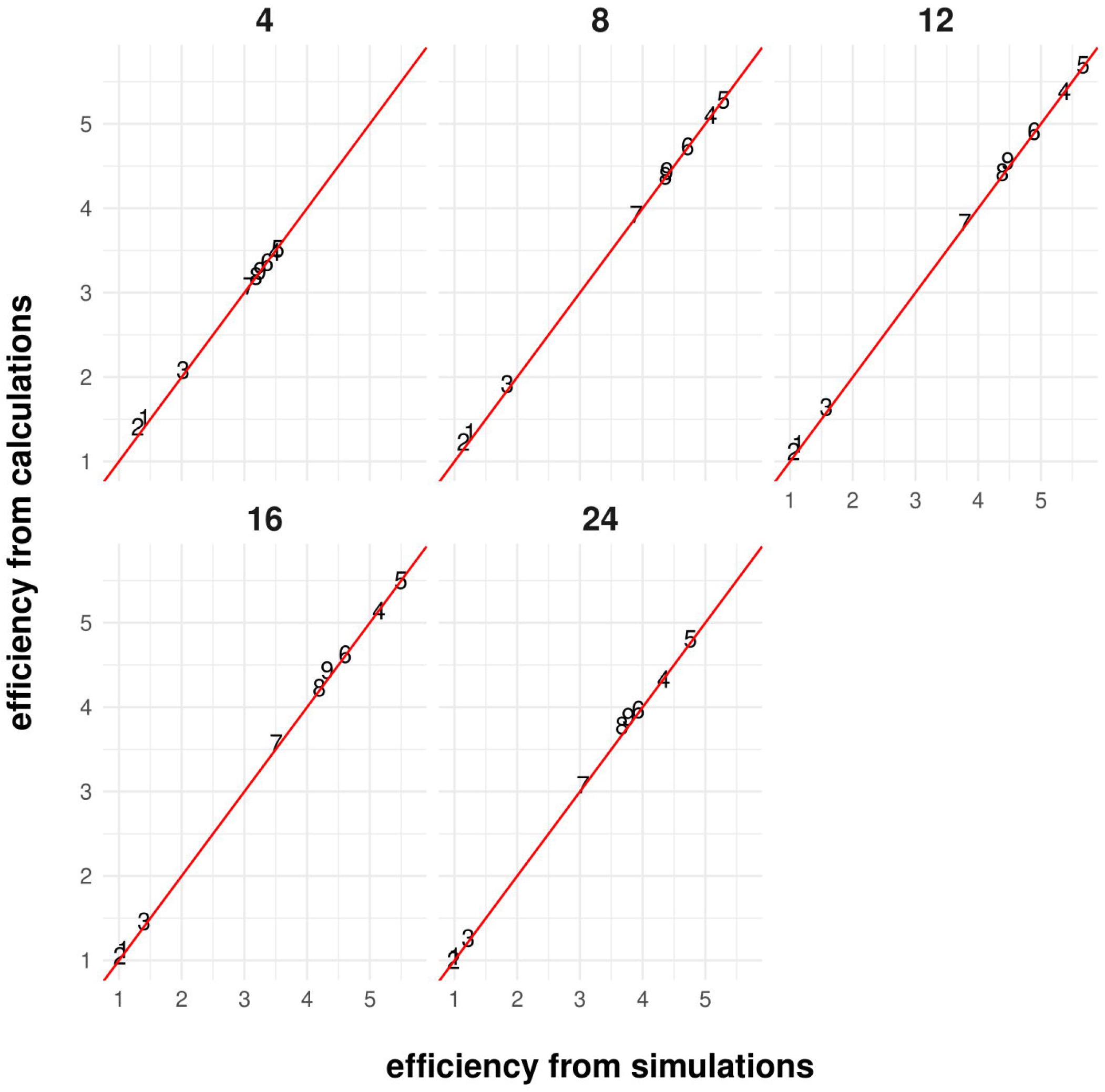

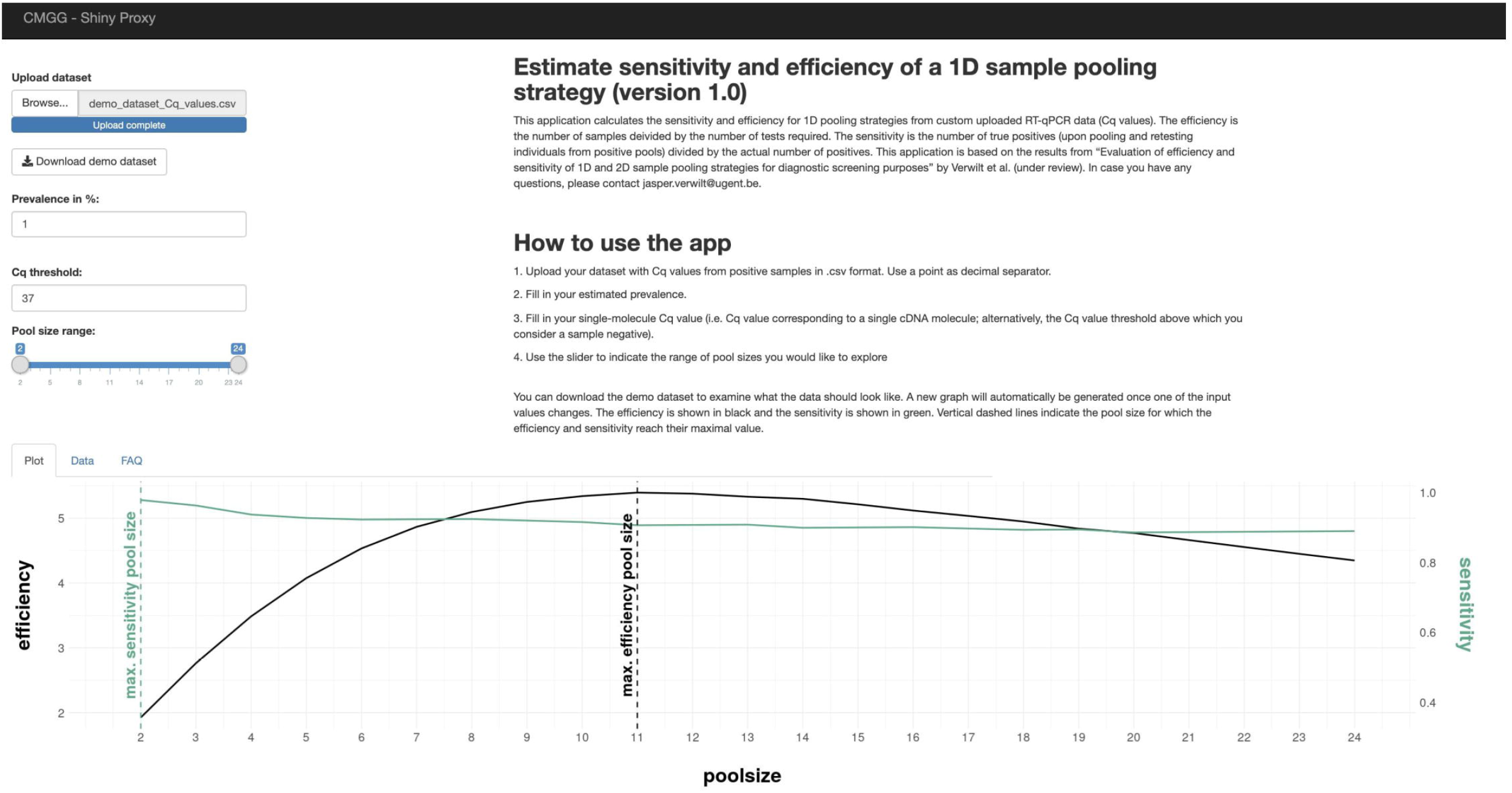

