## Supplemental Materials for "Evaluation of efficiency and sensitivity of 1D and 2D sample pooling strategies for SARS-CoV-2 RT-qPCR screening purposes"

- 5 supplemental figures

**Supplemental Figures**

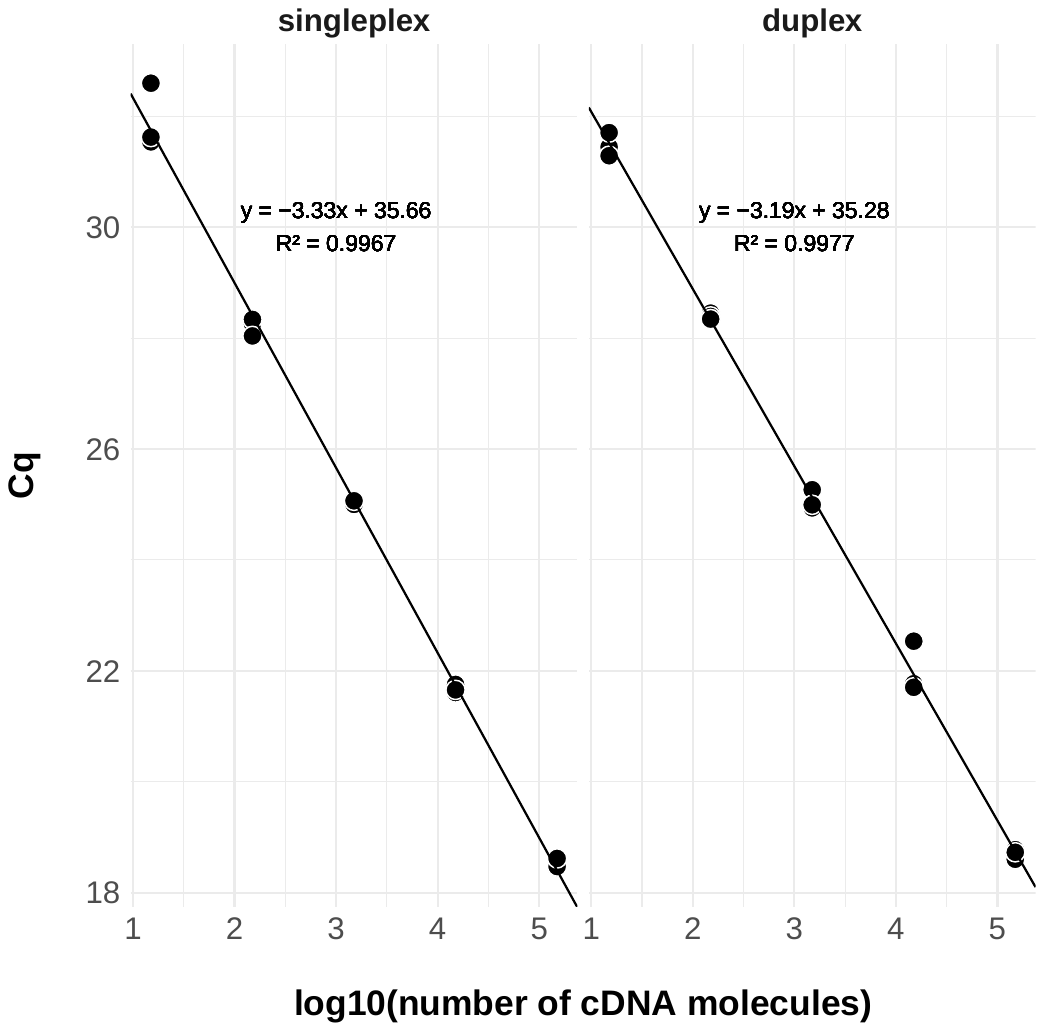

**Supplemental Figure 1:** Sensitivity analysis of singleplex and duplex qPCR assays using predetermined number of cDNA molecules. R-squared values are adjusted using the Wherry formula. Each number of cDNA molecules was tested in triplicate.

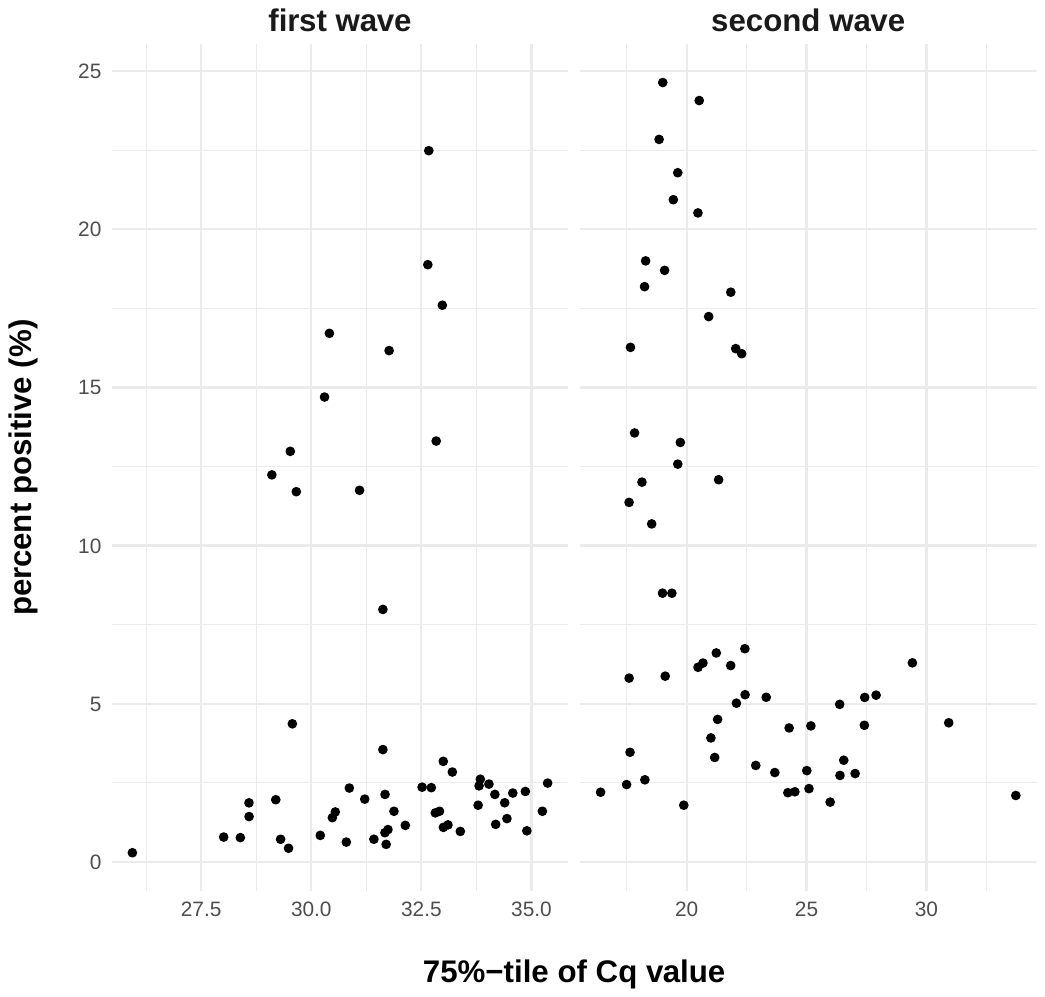

**Supplemental Figure 2:** Relationship of percent positive samples and 75%-tile of the Cq values. Each point is a specific day.

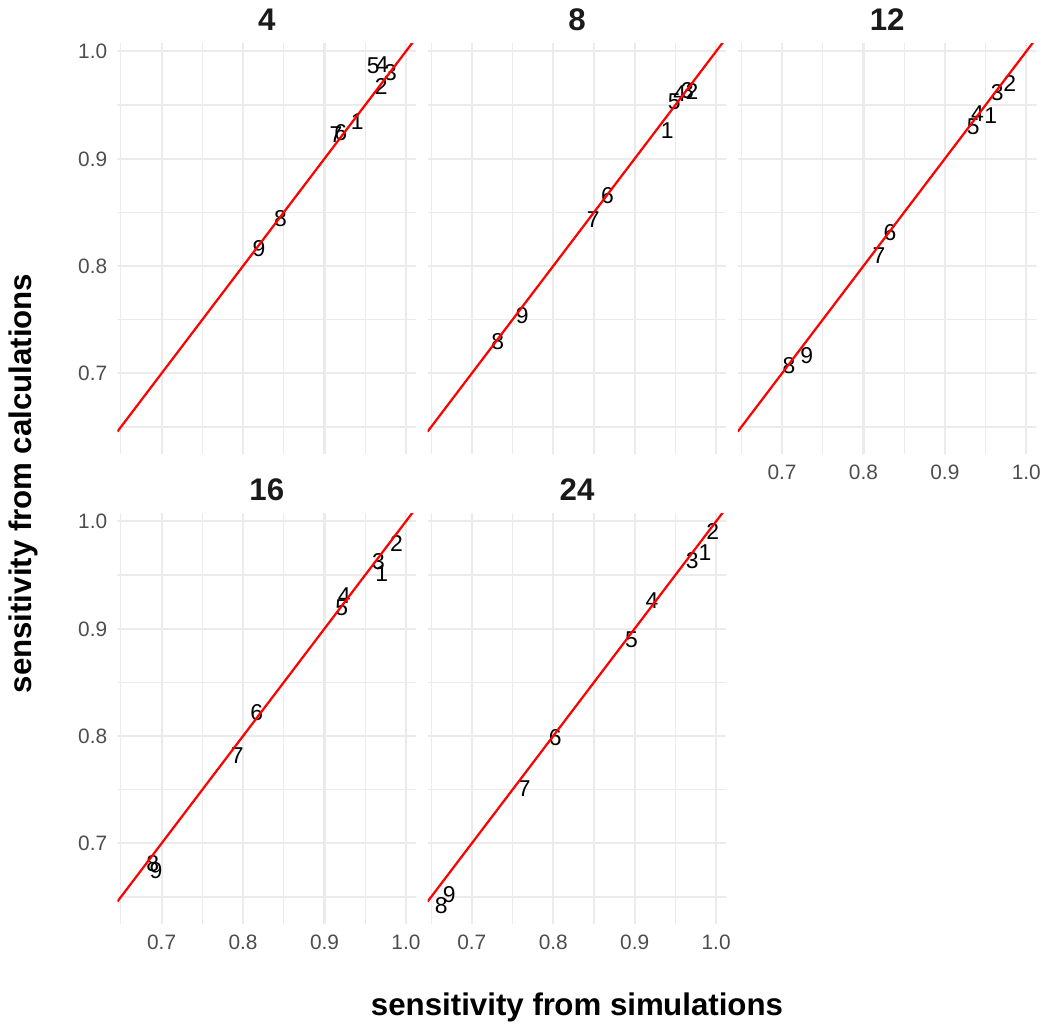

**Supplemental Figure 3:** Concordance of sensitivity estimations between simulations and calculations for the end of the first Belgian SARS-CoV-2 infection wave. The numbers represent the weeks (1: 1^st^ week; 2: 2^nd^ week; …) and are plotted at the sensitivities derived from the simulations and calculations. The red line represents the points where both values are equal.

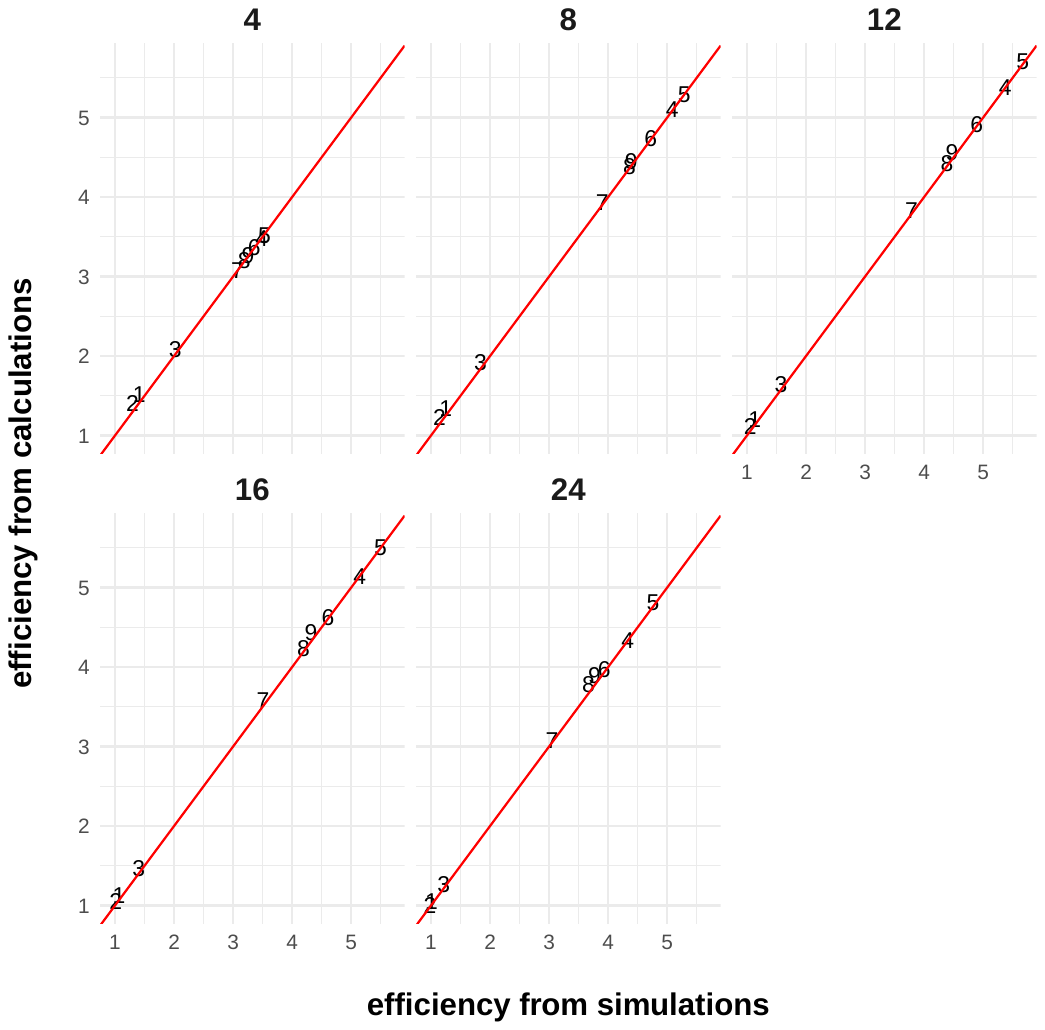

**Supplemental Figure 4:** Concordance of efficiency estimations between simulations and calculations for the end of the first Belgian SARS-CoV-2 infection wave. The numbers represent the weeks (1: 1^st^ week; 2: 2^nd^ week; …) and are plotted at the efficiencies derived from the simulations and calculations. The red line represents the points where both values are equal.

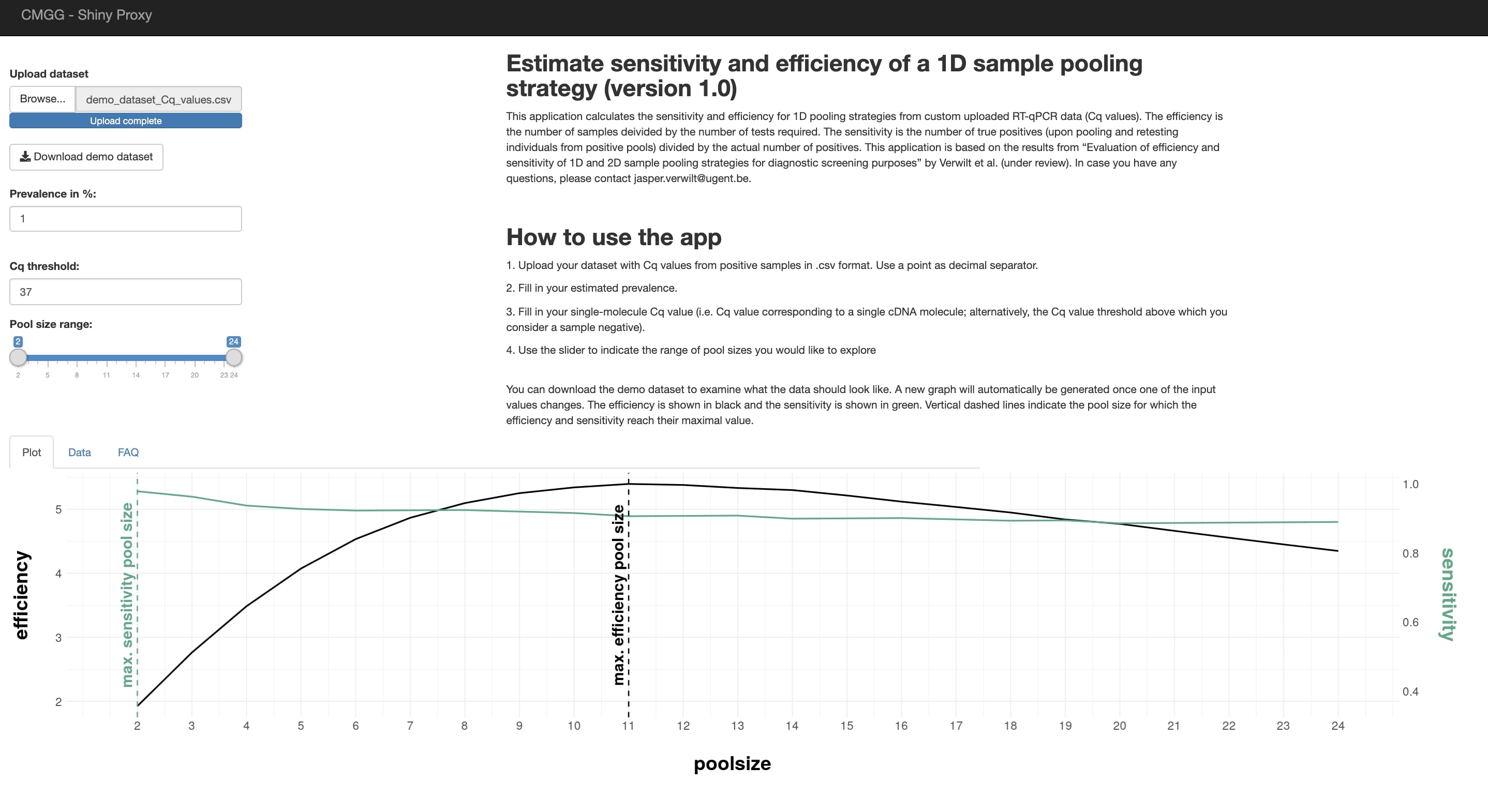

**Supplemental Figure 5:** A screenshot of the interface of the Shiny application. The webpage provides the user with a short description and a detailed outline of how to use the application. In the upper left corner, the user can provide their dataset. If the user would prefer to first explore the app without using their own data, a demo dataset can be downloaded and used instead. The user can fill in the estimated prevalence and single-molecule Cq value. The slider underneath can be used to indicated which range of pool sizes the user wishes to explore. Upon uploading the data, a graph will be outputted in the “Plot” tab, showing the estimated sensitivity and efficiency of each pool size. The vertical dashed lines represent the pool size at which the corresponding parameter reaches its maximal value for this data. The “Data” tab provides the user with a tabulated overview of the estimated sensitivity and efficiency of each pool size.
